# Association Between Angiotensin Receptor-Neprilysin Inhibitor Use and Clinical Outcomes in Concurrent Heart Failure with Reduced Ejection Fraction and End-Stage Renal Disease

**DOI:** 10.1101/2024.10.11.24315361

**Authors:** Mi-Hyang Jung, Dong-Hyuk Cho, Jimi Choi, Mi-Na Kim, Chan Joo Lee, Jung-Woo Son, Jong-Chan Youn, Byung-Su Yoo

## Abstract

**Background:** Although angiotensin receptor-neprilysin inhibitor (ARNI) therapy has been shown to improve outcomes in heart failure with reduced ejection fraction (HFrEF), its benefits in patients with end-stage renal disease (ESRD) on dialysis remain uncertain. This study investigated the clinical outcomes of ARNI compared to renin-angiotensin system (RAS) blockers in HFrEF patients with concomitant ESRD on dialysis.

**Methods:** Using the Korean National Health Insurance Service database, we identified individuals with HFrEF and ESRD on dialysis who were prescribed either ARNI or RAS blockers between 2017 and 2021. After applying inverse probability of treatment weighting, we compared 2,104 patients on ARNI with 2,191 on RAS blockers. The primary endpoint was a composite of all-cause mortality and any hospitalization over 2 years.

**Results:** Baseline characteristics were balanced between the groups. ARNI use was associated with a significantly lower risk of the primary endpoint (hazard ratio [HR] 0.86, 95% confidence interval [CI] 0.75–0.97) compared to RAS blockers. Additionally, ARNI was associated with a lower risk of all-cause mortality (HR 0.68, 95% CI 0.54–0.86) and any hospitalization (HR 0.86, 95% CI 0.75–0.98). Subgroup analyses demonstrated consistent associations between ARNI use and reduced risk across all subgroups (age, sex, comorbidities, and medications). Good adherence to ARNI was associated with a lower risk of the primary outcome, whereas non-adherence showed no such benefit.

**Conclusion:** In a real-world population with HFrEF and ESRD on dialysis, ARNI use was associated with a lower risk of all-cause mortality and hospitalization compared to RAS blockers, particularly in those with good adherence to therapy.

Graphical Abstract. Association between Angiotensin Receptor-Neprilysin Inhibitor Use and Clinical Outcomes in Patients with Concurrent Heart Failure with Reduced Ejection Fraction and End-Stage Renal Disease on Dialysis
In this real-world study of patients with heart failure with reduced ejection fraction (HFrEF) and end-stage renal disease (ESRD) on dialysis, the use of angiotensin receptor-neprilysin inhibitor (ARNI) was associated with a significant reduction in the risk of all-cause mortality and any hospitalization compared to renin-angiotensin system (RAS) blockers. These benefits were particularly evident in patients who adhered well to their medication.

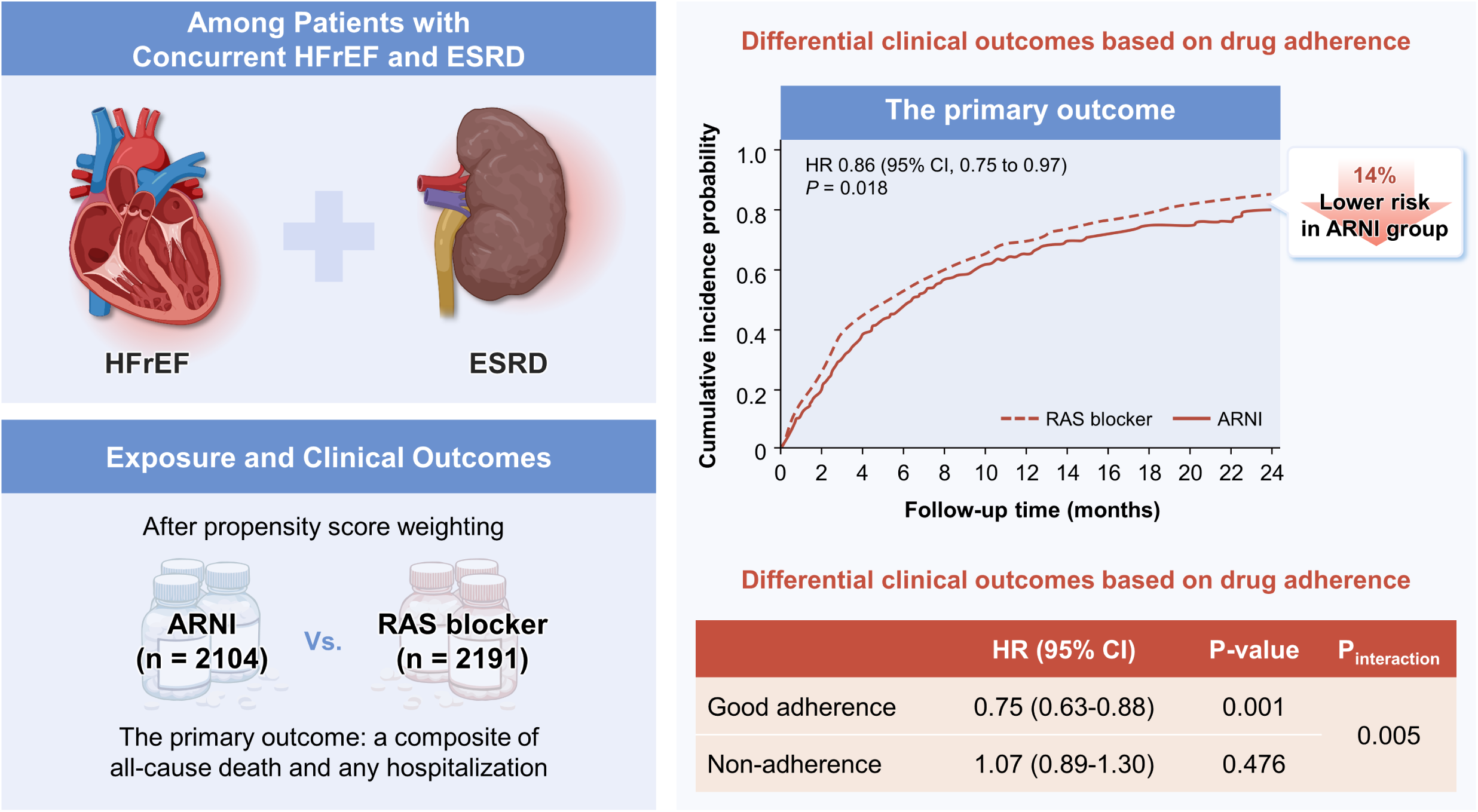

## Introduction

Patients with heart failure often have coexisting chronic kidney disease (CKD), with prevalence ranging from 26% to 64%, and approximately 5-6% of heart failure patients are on dialysis.^1–4^ In heart failure, deteriorating kidney function is associated with increased mortality, and this risk is even more pronounced in patients with end-stage renal disease (ESRD).^1,4,5^ Unfortunately, most medications for heart failure with reduced ejection fraction (HFrEF) lack strong evidence in patients with estimated glomerular filtration rate (eGFR) < 30 mL/min/1.73 m^2^, particularly those with ESRD.^4,6,7^ This underscores significant risks for patients with HFrEF and ESRD, highlighting unmet needs in treatment.

The angiotensin-receptor neprilysin inhibitor (ARNI) is considered a foundational medication for HFrEF due to its efficacy in reducing all-cause mortality and heart failure hospitalization, thereby forming one of the four pillars of treatment.^8,9^ However, patients with an eGFR < 30 mL/min/1.73 m² were excluded from the trial, and its efficacy in ESRD patients is not clear. A previous study by Lee et al. showed that ARNI improved left ventricular ejection fraction and reduced levels of soluble ST2 among patients with concurrent HFrEF and ESRD.^10^ Another recent case-control study demonstrated that ARNI use was associated with improved left ventricular systolic and diastolic function over one year, while these changes were not seen in the conventional treatment group among patients with concomitant HFrEF and ESRD.^11^ They also found that predialysis serum potassium levels did not significantly increase after ARNI prescription. However, these studies did not explore robust clinical outcomes, such as mortality or hospitalization.

Therefore, we conducted this real-world cohort study to elucidate the association between ARNI use and clinical outcomes compared to conventional treatment in patients with combined HFrEF and ESRD on dialysis. Furthermore, we aimed to examine whether adherence to the medication differentially affect the clinical outcomes in this population.

## Methods

### Data source and study population

This study utilized data from the Korean National Health Insurance System (NHIS), a comprehensive and mandatory health insurance program managed by the Korean government. The NHIS provides healthcare coverage to nearly the entire population of South Korea, ensuring a vast and inclusive database. This system operates on a fee-for-service model, enabling the collection of detailed medical expense claims for all insured individuals nationwide.^12–14^ Specifically, the NHIS database includes: (1) a qualification database, which contains basic information about enrollees such as age and sex; (2) a claims database, which includes diagnostic information classified by the International Classification of Diseases, Tenth Revision (ICD-10), and records of prescriptions and procedures for both inpatient and outpatient services; and (3) mortality data, which are linked to the database of Statistics Korea using unique identification numbers.

The eligible study population included individuals aged 18 years or older diagnosed with HFrEF who were taking either ARNI or traditional renin-angiotensin system (RAS) blockers (angiotensin-converting enzyme inhibitors or angiotensin receptor blockers) while also undergoing dialysis for ESRD between July 2017 and December 2021. HFrEF diagnosis was confirmed using the following criteria: 1) initiation of ARNI, indicating no previous use of ARNI for at least three years before the index date, or 2) at least two claims for RAS blockers under the following ICD-10 codes for heart failure (heart failure with systolic dysfunction [I50.04], left ventricular failure [I50.1], dilated cardiomyopathy [I42.0], or ischemic cardiomyopathy [I25.5]), along with examination of natriuretic peptide or echocardiography within six months of the RAS blocker prescription. For reference, ARNI is approved only for HFrEF patients with a left ventricular ejection fraction of ≤40% in Korea.

The Health Insurance Review and Assessment Service strictly evaluates ARNI prescriptions, making it a reliable indicator for HFrEF diagnosis. We excluded individuals who were diagnosed with cancer within the past 5 years, had undergone heart transplantation or left ventricular assist device implantation, had a human immunodeficiency virus infection, were admitted to a nursing hospital at the index date, had a total prescription duration for ARNI or RAS blockers of less than 90 days, or had a follow-up duration of less than 6 months. We further excluded those who only undergo continuous renal replacement therapy. The final cohort comprised 853 individuals on ARNI and 1,389 individuals on RAS blockers. After applying inverse probability treatment weighting (IPTW) using the propensity score for each subject, the final analysis included a weighted sample of 2,104 individuals on ARNI and 2,191 individuals on RAS blockers. The study flowchart is depicted in **supplementary Figure S1**.

### Study outcomes

The primary outcome was a composite of all-cause mortality and any hospitalization within 2 years. Secondary outcomes included all-cause mortality, any hospitalization, or cardiovascular mortality within 2 years. Cardiovascular mortality was defined as death occurring within 30 days after a diagnosis of the following diseases: myocardial infarction (I21-I23), unstable angina (I20.0), heart failure (I11.0, I13.0, I13.2, I50), cardiomyopathy (I42), stroke (I60-I64), sudden cardiac arrest (I46.9), cardiogenic shock (R57.0), other cerebrovascular events (I65-I69), and other cardiovascular events (I24-25, I30-49, I51-52). The index date was defined as the first prescription of either ARNI or a RAS blocker during the study period. From the index date, patients were monitored for up to two years or until the outcome occurred, following a 90-day supply of medication.

### Covariates

As covariates, we collected data on the cohort entry year, age, sex, socioeconomic status, region, duration of RAS blocker prescription before the index date, comorbidity burden, medication, and healthcare utilization. The cohort entry year was divided based on the COVID-19 period (pre-COVID-19 and amid-COVID-19), considering the potential impact of COVID-19 on clinical outcomes. Socioeconomic status was categorized into three groups (low, middle, and high) based on insurance premiums. The Charlson Comorbidity Index was calculated to assess the comorbidity burden.^15^

### Statistical analyses

Continuous variables were expressed as mean ± standard deviation or median (interquartile range), while categorical variables were presented as number (percentage). To reduce confounding, we adopted the IPTW method. The goal of IPTW is to estimate treatment causal effects more accurately by creating a weighted sample where the distribution of confounding variables or prognostically important covariates is approximately equal between the comparison groups.^16^ A multiple logistic regression model was used to estimate the propensity score, with the treatment group as the dependent variable and all baseline characteristics presented in Table 1 as independent variables. Baseline characteristics were summarized descriptively both before and after applying IPTW to evaluate comparability. Covariates were considered well-balanced if the absolute standardized difference was less than 0.1. Incidence rates were calculated as the weighted number of events by the weighted total person-time at risk and were presented as events per 100 person-years. The cumulative incidence of outcomes was graphically presented using a weighted Kaplan–Meier curve and compared between groups.^17^ The relationship between treatments and clinical outcomes was assessed using Cox proportional hazards regression with a robust sandwich-type variance estimator to account for the weights.^16^ The results were expressed as hazard ratios and 95% confidence intervals. The proportional hazards assumption of the Cox regression was tested based on Schoenfeld residuals. Subgroup analyses were conducted based on cohort entry year, age, sex, socioeconomic status, region, Charlson Comorbidity Index (divided into two groups based on its median value in our cohort), presence of diabetes, hypertension, ischemic heart disease, cerebrovascular disease, atrial fibrillation, and use of β-blockers and aldosterone antagonists. The results were presented as forest plots. Additionally, we analyzed the potential differences in clinical outcomes based on drug adherence. Drug adherence was evaluated using the proportion of days covered (PDC), which measures the percentage of days a patient has access to their prescribed medications over a specific period. A cutoff value of 80% was used to define adequate adherence, with good adherence defined as PDC ≥ 80% and non-adherence as PDC < 80%.^13^ Furthermore, we performed the same analyses using a cutoff value of 90%. All statistical analyses were performed using SAS Enterprise Guide software, version 7.1 (SAS Institute Inc., Cary, NC, USA) and R software, version 4.1 (R Foundation for Statistical Computing, Vienna, Austria), with a two-sided *P*-value of < 0.05 set as the threshold for statistical significance.

**Table 1.** Study population characteristics.

|  | Before IPTW |  |  | After IPTW |  |  |
| --- | --- | --- | --- | --- | --- | --- |
|  | ARNI<br>(n = 853) | RAS blocker<br>(n = 1,389) | ASD | ARNI<br>(Weighted n = 2,104) | RAS blocker<br>(Weighted n = 2,191) | ASD |
| **Cohort entry year** |  |  | 0.772 |  |  | 0.037 |
| 2017-2019 (Pre-COVID-19) | 338 (39.6) | 1,045 (75.2) |  | 1258 (59.8) | 1270 (58.0) |  |
| 2020-2021 (Amid COVID-19) | 515 (60.4) | 344 (24.8) |  | 846 (40.2) | 921 (42.0) |  |
| **Age** | 62.8 ± 12.3 | 63.1 ± 13.7 | 0.029 | 62.3 ± 19.9 | 62.8 ± 17.6 | 0.030 |
| **Sex** |  |  | 0.137 |  |  | 0.004 |
| Female | 235 (27.5) | 470 (33.8) |  | 628 (29.8) | 657 (30.0) |  |
| Male | 618 (72.5) | 919 (66.2) |  | 1476 (70.2) | 1534 (70.0) |  |
| **Socioeconomic status** |  |  | 0.047 |  |  | 0.045 |
| Low | 367 (43.0) | 571 (41.1) |  | 873 (41.5) | 916 (41.8) |  |
| Middle | 231 (27.1) | 376 (27.1) |  | 614 (29.2) | 599 (27.3) |  |
| High | 255 (29.9) | 442 (31.8) |  | 617 (29.3) | 676 (30.9) |  |
| **Region** |  |  | 0.136 |  |  | <0.001 |
| Metropolitan | 151 (17.7) | 310 (22.3) |  | 418 (19.9) | 430 (19.6) |  |
| City | 231 (27.1) | 303 (21.8) |  | 521 (24.8) | 545 (24.9) |  |
| Rural | 471 (55.2) | 776 (55.9) |  | 1,166 (55.4) | 1,216 (55.5) |  |
| **Duration of RAS blocker before the index date, month** | 32.2 ± 41.8 | 36.2 ± 41.7 | 0.085 | 34.8 ± 68.8 | 35.0 ± 54.4 | 0.003 |
| **Comorbidities, n (%)** |  |  |  |  |  |  |
| Charlson comorbidity index | 7 (6-9) | 7 (6-9) | 0.018 | 7 (6-9) | 7 (6-9) | 0.006 |
| Diabetes mellitus | 266 (31.2) | 409 (29.4) | 0.038 | 587 (27.9) | 653 (29.8) | 0.042 |
| Hypertension | 582 (68.2) | 955 (68.8) | 0.011 | 1,443 (68.6) | 1,505 (68.7) | 0.002 |
| Dyslipidemia | 393 (46.1) | 592 (42.6) | 0.070 | 915 (43.5) | 967 (44.1) | 0.012 |
| Pacemaker | 6 (0.7) | 9 (0.6) | 0.007 | 11 (0.5) | 14 (0.6) | 0.012 |
| ICD | 7 (0.8) | 7 (0.5) | 0.039 | 12 (0.6) | 12 (0.5) | 0.004 |
| CRT | 4 (0.5) | 5 (0.4) | 0.017 | 8 (0.4) | 18 (0.8) | 0.059 |
| Coronary revascularization | 176 (20.6) | 241 (17.4) | 0.084 | 404 (19.2) | 413 (18.8) | 0.009 |
| Ischemic heart disease | 612 (71.7) | 970 (69.8) | 0.042 | 1,428 (67.9) | 1,538 (70.2) | 0.050 |
| Angina | 513 (60.1) | 785 (56.5) | 0.074 | 1,192 (56.7) | 1,281 (58.5) | 0.037 |
| Myocardial infarction | 144 (16.9) | 179 (12.9) | 0.112 | 299 (14.2) | 315 (14.4) | 0.006 |
| Other ischemic heart disease | 333 (39.0) | 590 (42.5) | 0.070 | 810 (38.5) | 877 (40.0) | 0.031 |
| Cerebrovascular disease | 92 (10.8) | 184 (13.2) | 0.076 | 247 (11.7) | 264 (12.0) | 0.010 |
| Ischemic stroke | 76 (8.9) | 157 (11.3) | 0.080 | 206 (9.8) | 224 (10.2) | 0.013 |
| Hemorrhagic stroke | 12 (1.4) | 20 (1.4) | 0.003 | 34 (1.6) | 31 (1.4) | 0.015 |
| Transient ischemic attack | 12 (1.4) | 20 (1.4) | 0.003 | 27 (1.3) | 29 (1.3) | 0.006 |
| Peripheral artery disease | 57 (6.7) | 93 (6.7) | 0.001 | 149 (7.1) | 146 (6.7) | 0.015 |
| Thromboembolism | 40 (4.7) | 56 (4.0) | 0.032 | 91 (4.3) | 108 (4.9) | 0.029 |
| Atrial fibrillation | 152 (17.8) | 199 (14.3) | 0.095 | 357 (17.0) | 370 (16.9) | 0.003 |
| Sick sinus syndrome | 2 (0.2) | 9 (0.6) | 0.062 | 8 (0.4) | 10 (0.5) | 0.007 |
| **Medications, n (%)** |  |  |  |  |  |  |
| Diabetes medications |  |  |  |  |  |  |
| Metformin | 42 (4.9) | 66 (4.8) | 0.008 | 95 (4.5) | 97 (4.4) | 0.005 |
| Sulfonylurea | 208 (24.4) | 332 (23.9) | 0.011 | 509 (24.2) | 512 (23.4) | 0.020 |
| DPP-4 inhibitor | 377 (44.2) | 570 (41.0) | 0.064 | 886 (42.1) | 937 (42.8) | 0.013 |
| Thiazolidinedione | 16 (1.9) | 36 (2.6) | 0.049 | 57 (2.7) | 46 (2.1) | 0.040 |
| GLP1-RA | 13 (1.5) | 4 (0.3) | 0.131 | 17 (0.8) | 18 (0.8) | 0.005 |
| Insulin | 243 (28.5) | 410 (29.5) | 0.023 | 599 (28.5) | 644 (29.4) | 0.020 |
| Hypertension medications |  |  |  |  |  |  |
| Calcium channel blocker | 450 (52.8) | 938 (67.5) | 0.305 | 1,253 (59.6) | 1,306 (59.6) | 0.001 |
| Diuretics | 522 (61.2) | 763 (54.9) | 0.127 | 1,204 (57.2) | 1,251 (57.1) | 0.003 |
| Beta-blocker | 601 (70.5) | 935 (67.3) | 0.068 | 1,440 (68.4) | 1,515 (69.1) | 0.015 |
| Alpha-blocker | 148 (17.4) | 265 (19.1) | 0.045 | 328 (15.6) | 388 (17.7) | 0.057 |
| Digoxin | 36 (4.2) | 70 (5.0) | 0.039 | 89 (4.2) | 95 (4.3) | 0.004 |
| Nitrate | 140 (16.4) | 248 (17.9) | 0.038 | 327 (15.5) | 369 (16.8) | 0.036 |
| Lipid lowering agents |  |  |  |  |  |  |
| Statin | 506 (59.3) | 791 (56.9) | 0.048 | 1,199 (57.0) | 1,250 (57.1) | 0.001 |
| Others | 94 (11.0) | 138 (9.9) | 0.035 | 241 (11.5) | 237 (10.8) | 0.021 |
| Anticoagulants | 99 (11.6) | 118 (8.5) | 0.104 | 220 (10.5) | 225 (10.3) | 0.007 |
| Antiplatelet agent | 661 (77.5) | 1,037 (74.7) | 0.066 | 1,577 (75.0) | 1,656 (75.6) | 0.015 |
| **Healthcare Utilization** |  |  |  |  |  |  |
| No. of outpatient clinic visits, median (IQR) | 102 (33-169) | 86 (32-168) | 0.048 | 104 (31-170) | 95 (33-169) | 0.008 |
| Hospitalization within prior 1 year | 745 (87.3) | 1,135 (81.7) | 0.156 | 1,760 (83.7) | 1,836 (85.0) | 0.037 |
| ER visit | 25 (2.9) | 35 (2.5) | 0.025 | 47 (2.3) | 66 (3.0) | 0.047 |

ARNI, angiotensin receptor-neprilysin inhibitor; ASD, absolute standardized difference; DPP-4 inhibitor, dipeptidyl peptidase-4 inhibitor; ER, emergency room; GLP1-RA, glucagon-like peptide 1 receptor agonist; ICD, implantable cardioverter-defibrillator; IPTW, inverse probability treatment weighting; IQR, interquartile range; RAS, renin-angiotensin system.
Descriptive data were expressed as mean ± standard deviation or median (interquartile range).

### Ethics and data availability statement

The local institutional review board reviewed and approved the study protocol (IRB No. CR321358). Participant consent was waived because anonymized data were provided by the NHIS under a strict confidentiality protocol. The study findings are supported by data from the NHIS, which can be accessed upon review and approval of a study proposal by the NHIS.

## RESULTS

### Baseline demographic findings

After applying IPTW, baseline characteristics were well balanced between the groups (**Table 1**). We compared 2,104 patients on ARNI with 2,191 on RAS blockers. Overall, the mean age was 62.5 ± 18.8 years, with males accounting for 70% of the study population. In our cohort, the Charlson Comorbidity Index was relatively high, with a median (interquartile range) value of 7 (6-9).

### Association between ARNI use and clinical outcomes

During a median follow-up of 19.1 months (interquartile range 9.5–24.0), ARNI use was associated with a 14% lower risk of the primary outcome compared to the use of RAS blockers at 2 year (hazard ratio 0.86, 95% confidence interval 0.75-0.97, *P* = 0.018, **Table 2**). Specifically, ARNI use was linked to a lower risk of any hospitalization (hazard ratio 0.86, 95% confidence interval 0.75-0.98, *P* = 0.021) and all-cause mortality (hazard ratio 0.68, 95% confidence interval 0.54-0.86, *P* = 0.001). Additionally, ARNI use was associated with a decreased risk of cardiovascular mortality (hazard ratio 0.68, 95% confidence interval 0.52-0.89, *P* = 0.004). In parallel with these findings, Kaplan-Meier curves also showed a lower risk of adverse outcomes in the ARNI group compared to the RAS blocker group (**Figure 1**).

**Figure 1.**
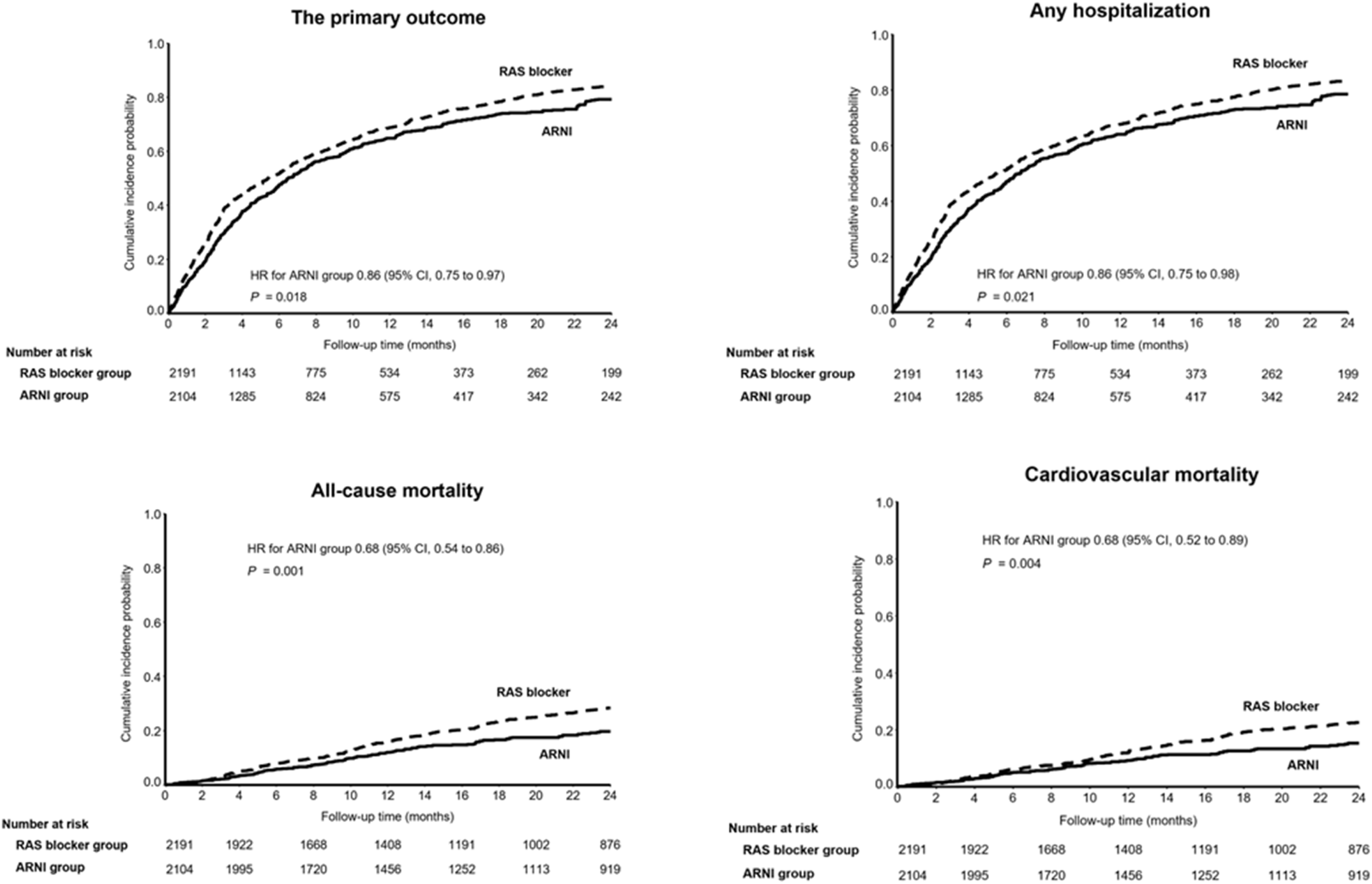
Weighted Kaplan-Meier curves for the primary and secondary outcomes. After IPTW, individuals on ARNI demonstrated a decreased risk of the primary outcome (a composite of all-cause mortality and any hospitalization) and the secondary outcomes (individual all-cause mortality, any hospitalization, and cardiovascular mortality) compared to those on RAS blockers in patients with concomitant HFrEF and ESRD on dialysis. Abbreviations: ARNI, angiotensin receptor-neprilysin inhibitor; CI, confidence interval; ESRD, end-stage renal disease; HFrEF, heart failure with reduced ejection fraction; IPTW, inverse probability of treatment weighting; RAS, renin-angiotensin system.

**Table 2.** The primary and secondary outcomes in the propensity score-matched population at 2 years.

| Outcomes | RAS blocker<br>(Weighted n = 2,191) | ARNI<br>(Weighted n = 2,104) | *P* |
| --- | --- | --- | --- |
| **The primary outcome** |  |  |  |
| No. of event (incidence rate per 100 PY) | 1,648 (118.50) | 1,516 (99.33) |  |
| Hazard ratio (95% confidence interval) | 1 (reference) | 0.86 (0.75-0.97) | 0.018 |
| **The secondary outcome** |  |  |  |
| All-cause mortality |  |  |  |
| No. of event (incidence rate per 100 PY) | 485 (16.69) | 340 (11.29) |  |
| Hazard ratio (95% confidence interval) | 1 (reference) | 0.68 (0.54-0.86) | 0.001 |
| Any hospitalization |  |  |  |
| No. of event (incidence rate per 100 PY) | 1,604 (115.34) | 1480 (96.99) |  |
| Hazard ratio (95% confidence interval) | 1 (reference) | 0.86 (0.75-0.98) | 0.021 |
| Cardiovascular mortality |  |  |  |
| No. of event (incidence rate per 100 PY) | 372 (12.81) | 259 (8.59) |  |
| Hazard ratio (95% confidence interval) | 1 (reference) | 0.68 (0.52-0.89) | 0.004 |

ARNI, angiotensin receptor-neprilysin inhibitor; IPTW, inverse probability treatment weighting; RAS, renin-angiotensin system.

### Subgroup analyses

The ARNI group exhibited a lower risk for the primary outcome than the RAS blocker group across various subgroups, including age, sex, cohort entry year categorized by the COVID-19 period, socioeconomic status, region, comorbidity burden, underlying cardio-cerebrovascular disease, atrial fibrillation, and other HFrEF medications (*P* for interaction > 0.05, **Figure 2**). Similar patterns were observed for any hospitalization, all-cause mortality, and cardiovascular mortality regardless of its subgroups (**supplementary Figures S2-4)**.

**Figure 2.**
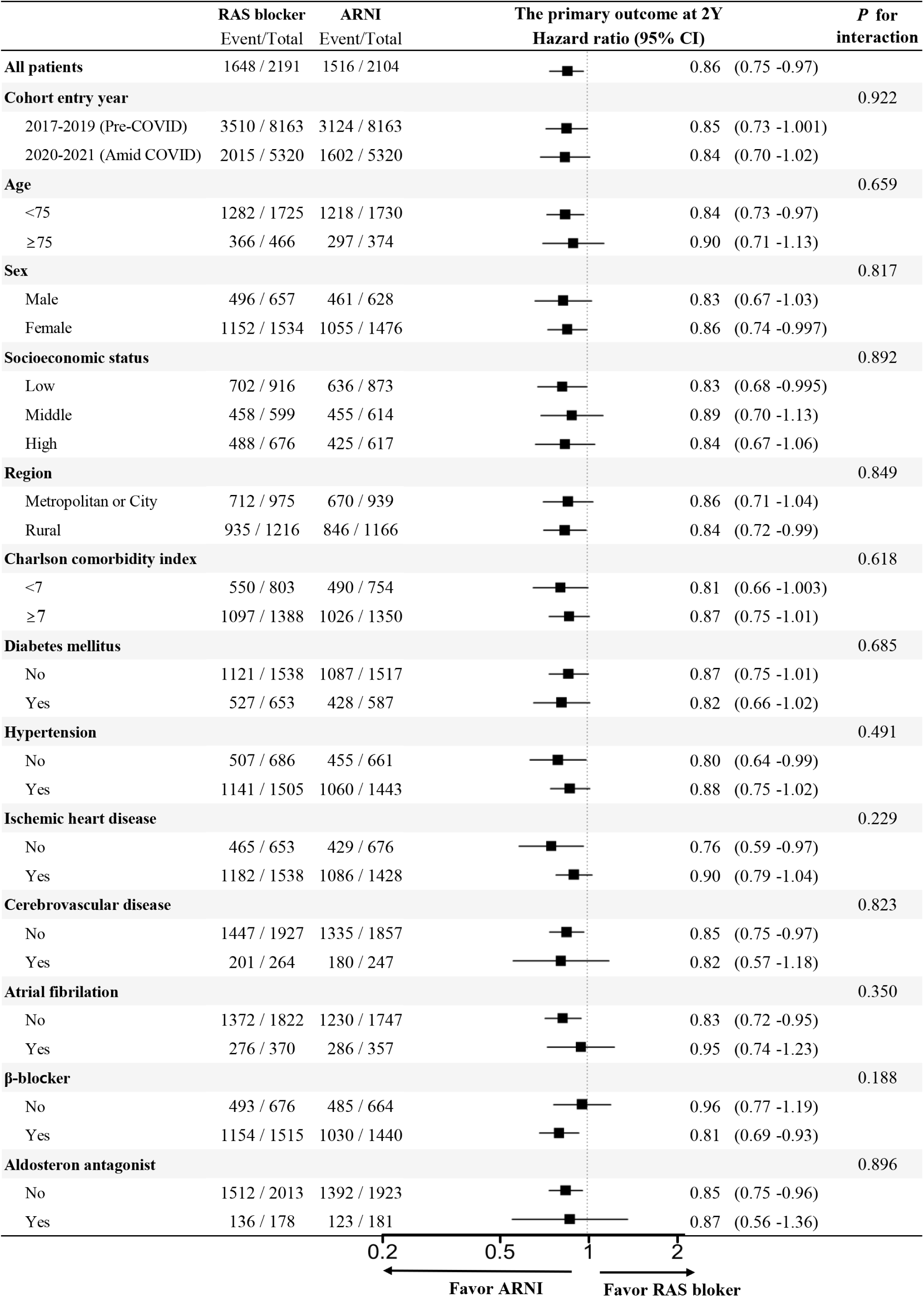
Subgroup analyses for the primary outcome. The forest plot displays hazard ratios for the primary outcome—a composite of all-cause mortality and any hospitalization at 2 years—comparing ARNI to RAS blockers across predefined subgroups in patients with HFrEF and ESRD on dialysis. Generally, ARNI is associated with a lower risk of adverse outcomes across various subgroups, with no significant interaction observed between treatment effects and the subgroups analyzed. Abbreviations: ARNI, angiotensin receptor-neprilysin inhibitor; CI, confidence interval; ESRD, end-stage renal disease; HFrEF, heart failure with reduced ejection fraction; RAS, renin-angiotensin system.

### Exploring clinical outcomes based on drug adherence

We explored whether differences in drug adherence might affect clinical outcomes. Good adherence to ARNI was clearly associated with a greater decrease in the primary outcome compared to good adherence to RAS blockers (hazard ratio 0.78, 95% confidence interval 0.67-0.90, *P* = 0.001). However, non-adherence to ARNI, defined as a PDC below 80%, did not show the same clinical benefit over non-adherence to RAS blockers (hazard ratio 1.00, 95% confidence interval 0.79-1.26, *P* = 0.995), although the *P*-value for interaction was 0.075 (**Table 3**). Particularly for all-cause mortality, there was a statistically significant interaction based on drug adherence (*P*-value for interaction 0.044), indicating that the decreased risk in all-cause mortality was only evident in the good adherence to ARNI group (**Table 3**).

**Table 3.** Clinical outcomes according to adherence to medication with a proportion of days covered (PDC) of 80%.

| Outcomes | RAS blocker (Ref.) |  | ARNI |  | Hazard ratio<br>(95% CI) | *P* | *P* for<br>interaction |
| --- | --- | --- | --- | --- | --- | --- | --- |
|  | Total | Event (IR) | Total | Event (IR) |  |  |  |
| The primary outcome |  |  |  |  |  |  |  |
| Good adherence | 1,468 | 1,299 (153.90) | 1,439 | 1,186 (115.19) | 0.78 (0.67-0.90) | 0.001 | 0.075 |
| Non-adherence | 723 | 349 (63.86) | 655 | 329 (66.38) | 1.00 (0.79-1.26) | 0.995 |  |
| The secondary outcome |  |  |  |  |  |  |  |
| All-cause mortality |  |  |  |  |  |  |  |
| Good adherence | 938 | 294 (20.48) | 1,077 | 202 (11.14) | 0.55 (0.40-0.74) | <0.001 | 0.044 |
| Non-adherence | 1,253 | 191 (13.00) | 1,027 | 138 (11.53) | 0.88 (0.62-1.25) | 0.474 |  |
| Any hospitalization |  |  |  |  |  |  |  |
| Good adherence | 1,468 | 1,266 (149.97) | 1,439 | 1,163 (112.91) | 0.79 (0.68-0.91) | 0.002 | 0.101 |
| Non-adherence | 723 | 338 (61.88) | 655 | 317 (63.90) | 0.99 (0.78-1.26) | 0.953 |  |
| Cardiovascular mortality |  |  |  |  |  |  |  |
| Good adherence | 938 | 232 (16.17) | 1,077 | 160 (8.82) | 0.55 (0.39-0.77) | 0.001 | 0.111 |
| Non-adherence | 1,253 | 140 (9.54) | 1,027 | 99 (8.23) | 0.86 (0.56-1.31) | 0.474 |  |

ARNI, angiotensin receptor-neprilysin inhibitor; IPTW, inverse probability treatment weighting; IR, event rate per 100 person year; RAS, renin-angiotensin system.
Drug adherence was defined by the proportion of days covered (PDC) for either a RAS blocker or ARNI, with a cutoff value of 80% (good adherence for PDC ≥ 80%, non-adherence for PDC $<$ 80%).

This trend became even more pronounced when the good adherence group was defined with a PDC cutoff of 90%, as illustrated in **Table 4**. Across all outcomes, there was a significant interaction between the good adherence group and the non-adherence group (*P*-value for interaction < 0.05). Good adherence to ARNI was associated with a significantly decreased risk in both the primary and secondary outcomes. In contrast, non-adherence (PDC < 90%) resulted in similar outcomes between the ARNI and RAS blocker groups.

**Table 4.** Clinical outcomes according to adherence to medication with a proportion of days covered (PDC) of 90%.

| Outcomes | RAS blocker (Ref.) |  | ARNI |  | Hazard ratio<br>(95% CI) | *P* | *P* for<br>interaction |
| --- | --- | --- | --- | --- | --- | --- | --- |
|  | Total | Event (IR) | Total | Event (IR) |  |  |  |
| The primary outcome |  |  |  |  |  |  |  |
| Good adherence | 1,281 | 1,145 (157.72) | 1,171 | 954 (111.28) | 0.75 (0.63-0.88) | 0.001 | 0.005 |
| Non-adherence | 910 | 503 (75.68) | 933 | 562 (84.01) | 1.07 (0.89-1.30) | 0.476 |  |
| The secondary outcome |  |  |  |  |  |  |  |
| All-cause mortality |  |  |  |  |  |  |  |
| Good adherence | 705 | 225 (20.56) | 825 | 145 (10.14) | 0.49 (0.34-0.71) | <0.001 | 0.023 |
| Non-adherence | 1,486 | 260 (14.36) | 1,279 | 190 (12.33) | 0.85 (0.63-1.15) | 0.300 |  |
| Any hospitalization |  |  |  |  |  |  |  |
| Good adherence | 1,281 | 1,123 (154.70) | 1,171 | 936 (109.14) | 0.75 (0.63-0.88) | 0.001 | 0.004 |
| Non-adherence | 910 | 481 (72.37) | 933 | 544 (81.40) | 1.08 (0.89-1.32) | 0.412 |  |
| Cardiovascular mortality |  |  |  |  |  |  |  |
| Good adherence | 705 | 178 (16.31) | 825 | 113 (7.94) | 0.49 (0.33-0.74) | 0.001 | 0.044 |
| Non-adherence | 1,486 | 194 (10.70) | 1,279 | 146 (9.17) | 0.85 (0.60-1.21) | 0.370 |  |

ARNI, angiotensin receptor-neprilysin inhibitor; IPTW, inverse probability treatment weighting; IR, event rate per 100 person year; RAS, renin-angiotensin system.
Drug adherence was defined by the proportion of days covered (PDC) for either a RAS blocker or ARNI, with a cutoff value of 90% (good adherence for PDC ≥ 90%, non-adherence for PDC < 90%).

## Discussion

In our real-world cohort of patients with concomitant HFrEF and ESRD on dialysis, ARNI was associated with a reduced risk of all-cause mortality and any hospitalization, underscoring its potential benefits in this under-evaluated population. This finding was consistent across various subgroups, including those defined by age, sex, cohort entry year, comorbidity burden, combined diseases, and other HFrEF medication use. Notably, good adherence to ARNI was linked to significant reductions in adverse clinical outcomes, while non-adherence did not confer the same benefits (**Graphical abstract**). Collectively, our data suggest a potential clinical benefit of ARNI, particularly emphasizing the importance of maintain adherence to optimize therapeutic outcomes.

Traditionally, individuals with ESRD have been in a therapeutic blind spot when it comes to HFrEF treatment. Despite being a high-risk population with significant mortality risk^1^ and challenges in volume control compared to the patients with preserved kidney function, they have had very few medications available, aside from RAS blockers and beta-blockers.^4,7^ Recently, ARNI has been established as a mainstay in HFrEF treatment after demonstrating its ability to reduce mortality and lower the risk of heart failure-related hospitalizations.^8,18,19^ Moreover, several studies have confirmed that ARNI provides the same benefits across a broader range of patients than those included in RCTs, with early initiation of ARNI demonstrating effectiveness in improving clinical outcomes and left ventricular remodeling.^20–24^ However, the effects of ARNI in patients with ESRD remain under-evaluated.

In this regard, our study provides valuable insight into this critical evidence gap. We found that patients prescribed ARNI had a significantly lower risk of the composite endpoint of all-cause mortality and any hospitalization compared to those receiving traditional treatment with RAS blockers. Notably, the clinical benefit of ARNI was only evident in patients who were highly adherent to their medication, emphasizing the crucial role of adherence in achieving the full benefits of ARNI. Supporting our findings, recent study demonstrated that ARNI significantly reduces cardiac biomarkers and improves left ventricular systolic function in patients with concomitant HFrEF and ESRD on dialysis.^10^ Specifically, high-sensitive troponin T levels decreased from 236.2 ± 355.3 to 97.0 ± 14.0 pg/mL, soluble ST2 levels dropped from 40.4 ± 44.0 to 19.6 ± 14.1 ng/mL, and left ventricular ejection fraction increased from 29.7 ± 4.4% to 40.8 ± 10.5% (all *P* < 0.05). These findings suggest that the benefits of reverse cardiac remodeling and reduced cardiac fibrosis through ARNI are applicable to patients with ESRD as well. Another study further corroborated these findings, reporting significant reverse cardiac remodeling in ARNI-treated patients with HFrEF and ESRD, an effect not observed in the conventional treatment group.^11^ After one year of ARNI treatment, left ventricular ejection fraction increased from 31.3% to 45.1%, left ventricular end-systolic volume decreased from 95.7 mL to 70.1 mL, the E/A ratio decreased from 1.3 to 0.8, and the medial E/e’ ratio improved from 25.3 to 18.8.^11^

A plausible explanation for ARNI’s favorable effects may lie in its dual action of augmenting the natriuretic peptide system and suppressing the over-activated renin-angiotensin-aldosterone system, mechanisms that are also relevant in patients with advanced CKD, including those with ESRD.^25^ By enhancing natriuretic peptides, ARNI improves cardiovascular and renal outcomes through several pathways, including promoting vasodilation, lowering blood pressure, reducing sympathetic nervous system activity, and increasing diuresis and natriuresis. Collectively, these effects reduce cardiac stress, prevent further remodeling, and protect renal function.^25^ The renal benefits of ARNI are especially significant in the context of the complex cardio-renal interaction.^26–28^ Previous research has demonstrated that ARNI slows the decline in glomerular filtration rate compared to enalapril, while consistently providing cardiovascular benefits in patients with or without CKD or albuminuria.^26^ Moreover, the more pronounced clinical benefits of ARNI in patients with good adherence, as shown in our study, further supports the efficacy of this medication in improving clinical outcomes.

Several limitations of this study should be acknowledged. First, although we applied IPTW for possible confounders, our study is based on a cohort design, which limits the ability to establish causality. Therefore, our findings should be considered hypothesis-generating, and further randomized clinical trials or large-scale cohort studies are necessary to confirm these results. Second, we lacked detailed echocardiographic and biomarker data, which could have provided deeper insights into the mechanisms involved. Third, we focused on any hospitalization instead of the more commonly used outcome of heart failure hospitalization because claim data may not reliably capture heart failure-specific hospitalizations. Therefore, we selected this broader outcome as a more robust and accurate measure. Additionally, we conducted subgroup analyses based on the cohort entry period during the COVID-19 pandemic to mitigate potential biases related to changes in hospitalization patterns during that time. Our results remained consistent regardless of the COVID-19 period. Finally, the follow-up period was relatively short (2 years), which may limit the long-term applicability of our findings. Despite these limitations, our study offers important observations by suggesting the potential benefit of ARNI in a key subgroup that has not been fully evaluated in randomized clinical trials. Additionally, it underscores the important role of medication adherence in optimizing the effectiveness of ARNI in real-world settings.

## Conclusions

In this real-world study of patients with HFrEF and ESRD on dialysis, ARNI was associated with a significant reduction in the risk of all-cause mortality and any hospitalization compared to RAS blockers. These benefits were particularly evident in patients who adhered well to their medication, highlighting the importance of adherence in maximizing therapeutic outcomes. Although our study is limited by its observational design and relatively short follow-up, the findings suggest that ARNI may offer a valuable treatment option for this high-risk population, warranting further investigation through randomized clinical trials.

## Data Availability

The study findings are supported by data from the NHIS, which can be accessed upon review and approval of a study proposal by the NHIS.

## Acknowledgments

The authors express their sincere gratitude to the personnel of Big Data Steering Department at NHIS for their valuable assistance and for providing the data.

## References

1. Damman K, Valente MA, Voors AA, O’Connor CM, van Veldhuisen DJ, Hillege HL. Renal impairment, worsening renal function, and outcome in patients with heart failure: an updated meta-analysis. Eur Heart J. 2014;35:455–469.

2. Hebert K, Dias A, Delgado MC, Franco E, Tamariz L, Steen D, et al. Epidemiology and survival of the five stages of chronic kidney disease in a systolic heart failure population. Eur J Heart Fail. 2010;12:861–865.

3. Patel RB, Fonarow GC, Greene SJ, Zhang S, Alhanti B, DeVore AD, et al. Kidney Function and Outcomes in Patients Hospitalized With Heart Failure. J Am Coll Cardiol. 2021;78:330343.

4. Khan MS, Ahmed A, Greene SJ, Fiuzat M, Kittleson MM, Butler J, et al. Managing Heart Failure in Patients on Dialysis: State-of-the-Art Review. J Card Fail. 2023;29:87–107.

5. Foley RN. Clinical epidemiology of cardiac disease in dialysis patients: left ventricular hypertrophy, ischemic heart disease, and cardiac failure. Semin Dial. 2003;16:111–117.

6. Beldhuis IE, Lam CSP, Testani JM, Voors AA, Van Spall HGC, Ter Maaten JM, et al. Evidence-Based Medical Therapy in Patients With Heart Failure With Reduced Ejection Fraction and Chronic Kidney Disease. Circulation. 2022;145:693–712.

7. Mullens W, Martens P, Testani JM, Tang WHW, Skouri H, Verbrugge FH, et al. Renal effects of guideline-directed medical therapies in heart failure: a consensus document from the Heart Failure Association of the European Society of Cardiology. Eur J Heart Fail. 2022;24:603–619.

8. McMurray JJ, Packer M, Desai AS, Gong J, Lefkowitz MP, Rizkala AR, et al. Angiotensin-neprilysin inhibition versus enalapril in heart failure. N Engl J Med. 2014;371:993–1004.

9. Youn JC, Kim D, Cho JY, Cho DH, Park SM, Jung MH, et al. Korean Society of Heart Failure Guidelines for the Management of Heart Failure: Treatment. Int J Heart Fail. 2023;5:66–81.

10. Lee S, Oh J, Kim H, Ha J, Chun KH, Lee CJ, et al. Sacubitril/valsartan in patients with heart failure with reduced ejection fraction with end-stage of renal disease. ESC Heart Fail. 2020;7:1125–1129.

11. Niu CY, Yang SF, Ou SM, Wu CH, Huang PH, Hung CL, et al. Sacubitril/Valsartan in Patients With Heart Failure and Concomitant End-Stage Kidney Disease. J Am Heart Assoc. 2022;11:e026407.

12. Lee CJ, Lee H, Yoon M, Chun KH, Kong MG, Jung MH, et al. Heart Failure Statistics 2024 Update: A Report From the Korean Society of Heart Failure. Int J Heart Fail. 2024;6:56–69.

13. Jung MH, Lee SY, Youn JC, Chung WB, Ihm SH, Kang D, et al. Antihypertensive Medication Adherence and Cardiovascular Outcomes in Patients With Cancer: A Nationwide Population-Based Cohort Study. J Am Heart Assoc. 2023;12:e029362.

14. Jung MH, Choi YS, Yi SW, An SJ, Yi JJ, Ihm SH, et al. Socioeconomic status and cardiovascular mortality in over 170,000 cancer survivors. Eur Heart J Qual Care Clin Outcomes. 10.1093/ehjqcco/qcae055.

15. Charlson ME, Pompei P, Ales KL, MacKenzie CR. A new method of classifying prognostic comorbidity in longitudinal studies: development and validation. J Chronic Dis. 1987;40:373–383.

16. Austin PC, Stuart EA. Moving towards best practice when using inverse probability of treatment weighting (IPTW) using the propensity score to estimate causal treatment effects in observational studies. Stat Med. 2015;34:3661–3679.

17. Xie J, Liu C. Adjusted Kaplan-Meier estimator and log-rank test with inverse probability of treatment weighting for survival data. Stat Med. 2005;24:3089–3110.

18. Velazquez EJ, Morrow DA, DeVore AD, Duffy CI, Ambrosy AP, McCague K, et al. Angiotensin-Neprilysin Inhibition in Acute Decompensated Heart Failure. N Engl J Med. 2019;380:539–548.

19. Wachter R, Senni M, Belohlavek J, Straburzynska-Migaj E, Witte KK, Kobalava Z, et al. Initiation of sacubitril/valsartan in haemodynamically stabilised heart failure patients in hospital or early after discharge: primary results of the randomised TRANSITION study. Eur J Heart Fail. 2019;21:998–1007.

20. Oh JH, Lee JM, Lee HJ, Hwang J, Lee CH, Cho YK, et al. The benefits of the earlier use of sacubitril/valsartan in de novo heart failure with reduced ejection fraction patients. ESC Heart Fail. 2022;9:2435–2444.

21. Januzzi JL Jr, Prescott MF, Butler J, Felker GM, Maisel AS, McCague K, et al. Association of Change in N-Terminal Pro-B-Type Natriuretic Peptide Following Initiation of Sacubitril-Valsartan Treatment With Cardiac Structure and Function in Patients With Heart Failure With Reduced Ejection Fraction. JAMA. 2019;322:1085–1095.

22. Park JJ, Lee SE, Cho HJ, Choi JO, Yoo BS, Kang SM, et al. Real-World Usage of Sacubitril/Valsartan in Korea: A Multi-Center, Retrospective Study. Int J Heart Fail. 2022;4:193–204.

23. Cho DH, Choi J, Youn JC, Yoo BS. The Comparative Effectiveness Between Angiotensin Receptor-neprilysin Inhibitor And Renin-angiotensin System Blockade In Patient With Heart Failure With Reduced Ejection Fraction: Nationwide Medication Adherence Study. J Card Fail. 2024;30:158. 10.1016/j.cardfail.2023.10.102.

24. Moon MG, Hwang IC, Lee HJ, Kim SH, Yoon YE, Park JB, et al. Reverse Remodeling Assessed by Left Atrial and Ventricular Strain Reflects Treatment Response to Sacubitril/Valsartan. JACC Cardiovasc Imaging. 2022;15:1525–1541.

25. Greenberg B. Angiotensin Receptor-Neprilysin Inhibition (ARNI) in Heart Failure. Int J Heart Fail. 2020;2:73–90.

26. Damman K, Gori M, Claggett B, Jhund PS, Senni M, Lefkowitz MP, et al. Renal Effects and Associated Outcomes During Angiotensin-Neprilysin Inhibition in Heart Failure. JACC Heart Fail. 2018;6:489–498.

27. Packer M, Claggett B, Lefkowitz MP, McMurray JJV, Rouleau JL, Solomon SD, et al. Effect of neprilysin inhibition on renal function in patients with type 2 diabetes and chronic heart failure who are receiving target doses of inhibitors of the renin-angiotensin system: a secondary analysis of the PARADIGM-HF trial. Lancet Diabetes Endocrinol. 2018;6:547–554.

28. Haynes R, Judge PK, Staplin N, Herrington WG, Storey BC, Bethel A, et al. Effects of Sacubitril/Valsartan Versus Irbesartan in Patients With Chronic Kidney Disease. Circulation. 2018;138:1505–1514.

